# EEG-based Machine Learning Models for the Prediction of Phenoconversion Time and Subtype in iRBD

**DOI:** 10.1101/2023.09.04.23294964

**Authors:** El Jeong, Yong Woo Shin, Jung-Ick Byun, Jun-Sang Sunwoo, Monica Roascio, Pietro Mattioli, Laura Giorgetti, Francesco Famà, Gabriele Arnulfo, Dario Arnaldi, Han-Joon Kim, Ki-Young Jung

## Abstract

**Background:** Idiopathic/Isolated rapid eye movement sleep behavior disorder (iRBD) is a prodromal stage of α-synucleinopathies and eventually phenoconverts to overt neurodegenerative diseases including Parkinson’s disease (PD), dementia with Lewy bodies (DLB) and multiple system atrophy (MSA). Associations of baseline resting-state electroencephalography (EEG) with phenoconversion have been reported.

**Objectives:** In this study, we aimed to develop machine learning models to predict phenoconversion time and subtype using baseline EEG features in patients with iRBD.

**Methods:** At baseline, resting-state EEG and neurological assessments were performed on patients with iRBD. Calculated EEG features included spectral power, weighted phase lag index and Shannon entropy. Three models were used for survival prediction, and four models were used for α-synucleinopathy subtype prediction. The models were externally validated using data from a different institution.

**Results:** A total of 236 iRBD patients were followed-up for up to eight years (mean 3.5 years), and 31 patients converted to α-synucleinopathies (16 PD, 9 DLB, 6 MSA). The best model for survival prediction was the random survival forest model with an integrated Brier score of 0.114 and a concordance index of 0.775. The K-nearest neighbor model was the best model for subtype prediction with an area under the receiver operating characteristic curve of 0.901. EEG slowing was an important feature for both models.

**Conclusions:** Machine learning models using baseline EEG features can be used to predict phenoconversion time and its subtype in patients with iRBD. Further research including large sample data from many countries is needed to make a more robust model.

## Introduction

Rapid eye movement (REM) sleep behavior disorder (RBD) is characterized by dream enactment and loss of atonia during REM sleep.^1^ Idiopathic/Isolated RBD (iRBD) is known as a prodromal stage of α-synucleinopathies, specifically Parkinson’s disease (PD), dementia with Lewy bodies (DLB) and multiple system atrophy (MSA).^2,3^ The risk of developing α-synucleinopathy among iRBD patients is approximately 18% after three years, 31% after five years and 74% after 12 years.^2^ In short, most iRBD patients eventually develop an α-synucleinopathy, preceded by a decline in either motor or cognitive function.^4^

Age, olfactory function, cognitive function and motor function have been reported as clinical biomarkers to predict phenoconversion with a hazard ratio up to 3.16.^5,6^ Neuroimaging of dopamine transporters provides a promising biomarker to predict phenoconversion;^7^ however, it is expensive and has limited accessibility. Electroencephalography (EEG) is a safe and easy method to objectively measure brain activity. EEG in iRBD patients previously showed slowing in the occipital region, decreased delta-band weighted phase lag index (wPLI) in the frontal regions, and increased alpha wPLI with reduced delta orthogonalized Correlation Coefficient (oCC).^8–11^ Two longitudinal studies have evaluated the value of EEG in predicting phenoconversion in iRBD.^12,13^ These studies, however, described phenoconversion dichotically, i.e., converted or not converted.

However, it is demanded for both patients and clinicians to predict ***when and how fast*** phenoconversion to α-synucleinopathy will occur and ***to which subtype*** of synucleinopathy the patient will phenoconvert. An individualized model that can predict the time from diagnosis until each patient develops a α-synucleinopathy is important in understanding their prognosis. Moreover, estimating whether the iRBD patients will first develop motor or cognitive symptoms is also crucial.

Machine learning is being extensively explored for potential applications in various diseases and has achieved excellent performance compared with conventional methods.^14^ Thus, machine learning methods can be considered for the application of predicting the complex survival time and subtype of iRBD phenoconversion. The aim of this study was to propose EEG-based machine learning models that can predict the time to phenoconversion and the subtype of phenoconversion for each patient. Various survival analyses and classification models were compared to select the best model.

## Methods

### Participants

Patients with iRBD who visited the sleep clinic of Seoul National University Hospital were enrolled and followed up every year. RBD was diagnosed according to the International Classification of Sleep Disorders - third edition (ICSD-3) criteria using overnight video-polysomnography (vPSG).^15^ Two neurologists specialized in sleep disorders (JK) and movement disorders (KH) examined each patient at baseline to evaluate them for dementia, cerebellar ataxia, parkinsonism or other neurodegenerative diseases.

Participants with a neurodegenerative disease, neurological disorder, severe medical illness or severe obstructive sleep apnea (apnea-hypopnea index ≥ 30) were excluded. This study was authorized by the Institutional Review Board (IRB) of the Seoul National University Hospital (IRB Number 1406-100-589). Written informed consent was obtained from each participant.

For external validation, we used clinical and EEG data of iRBD patients provided by the University Neurology Clinics at Policlinico San Martino in Genoa. Clinical and EEG data has been described in detail elsewhere.^11^ In brief, inclusion/exclusion criteria, clinical and EEG assessments substantially overlapped with the Korean cohort. All patients completed routine clinical follow-ups during which systematic assessments for parkinsonism and dementia were performed, including a semistructured interview with patients and caregivers (IRB Number 703, from the Genoa IRB).

### Clinical evaluation

The Korean version of the Mini-Mental Status Examination (K-MMSE) and the Korean version of the Montreal Cognitive Assessment (MoCA-K) were used to evaluate general cognitive function.^16,17^ The Korean version of the RBD Screening Questionnaire-Hong Kong (RBDQ-KR) was used to assess the RBD symptom severity.^18^ The Korean Version of Sniffing Sticks (KVSS) was applied to test olfactory symptoms.^19^ The Scales for Outcomes in Parkinson’s Disease for Autonomic Symptoms (SCOPA-AUT) questionnaire was used to examine the symptoms of autonomic dysfunction.^20^ The Movement Disorder Society — Unified Parkinson’s Disease Rating Scale (MDS-UPDRS) part III was used to assess motor symptoms.^21^ Additionally, subjective sleep quality and excessive daytime sleepiness were assessed using the Pittsburgh Sleep Quality Index (PSQI) and the Epworth Sleepiness Scale (ESS), respectively.^22,23^

During follow-up, cognitive function (K-MMSE, MoCA-K), motor function (MDS-UPDRS part III), autonomic function (SCOPA-AUT), self-reported sleep propensity and quality (PSQI, ESS), RBD symptom severity (RBDQ-KR) and olfactory function (KVSS) were evaluated every year. Phenoconversion in iRBD patients was assessed every 6 to 12 months by the same two neurologists (JK and KH). Finally, patients with iRBD who developed PD, DLB or MSA were classified as converters (iRBD-C), while the remaining patients were classified as nonconverters (iRBD-NC). The diagnoses of PD, DLB and MSA were made according to standard criteria.^24–26^

### EEG recordings and preprocessing

Scalp EEGs were obtained using a 60-channel EEG cap (Wave-Guard EEG cap, Advanced NeuroTechnology, Enschede, Netherlands) arranged according to the international 10-10 system. The reference electrode was positioned on an ear and the ground electrode was placed on the AFz. Impedances were kept under 10 kΩ. To detect and eliminate eye movement artifacts, two EOG electrodes were attached to the left and right outer canthi. The sampling rate was 400 Hz. The resting-state EEG was recorded for a total of 5 minutes in all patients while they were awake and alternating opening and closing their eyes every 30 seconds. To preprocess the data, a 0.5 Hz high-pass filter and a 60 Hz notch filter were applied. Only the EEG data recorded while the participant’s eyes were closed were extracted and analyzed in this study. EEG segments with severe artifacts or poor signal quality were removed by visually inspecting the data. Then, independent component analysis (ICA) was applied, and the EEGLAB plugin ICLabel was used to automatically remove eye artifacts.^27,28^ The threshold for eye artifact probability was set to 90%.

EEG data for the external validation set used a system with 61 electrodes according to the international 10-10 system. The reference and the ground electrode were Fpz and Oz, respectively, and the signals were sampled at 512 Hz. We simultaneously recording electrooculogram to monitor eye-movements. The acquisition protocol consisted of approximately 25 resting states recordings subdivided into 2–3 minutes with eyes open, 3–4 minutes during hyperventilation and 17–18 minutes with eyes closed. Impedances were kept under 5 kΩ. The same preprocessing procedures used for our dataset were implemented for the external validation set.

For both centers’ data, a total of 101 seconds of EEG data for each patient were eventually included in this study. EEG preprocessing was performed using the EEGLAB package (version 2019.1) for MATLAB (version 9.8.0, The MathWorks, Natick, MA, USA).^29^

### Experimental procedures

#### EEG features

To make the model more robust and reduce overfitting, data augmentation was performed for the training set. To augment the total data size the first 100 two-second EEG epochs were extracted by the sliding window method with 50% overlap. Thus, one patient’s EEG data was augmented to one hundred EEG epochs.

For each EEG epoch, the fast Fourier transforms (FFT) using the Hanning window was applied with a frequency of interest range of 1–50 Hz in 0.5 Hz steps. In our study, four frequency bands were used: delta (2-3.5 Hz), theta (4-7.5 Hz), alpha (8-12.5 Hz), beta (13-30 Hz). Absolute power was averaged across all electrodes and converted decibels. Relative power was calculated by expressing the percentage of power for each frequency band over the total power in the 2–30 Hz range. The dominant occipital frequency (DOF) was defined as the averaged peak frequency with the maximum power between 4–14 Hz in the two occipital channels (O1, O2). The slow-to-fast power ratio (STF) was calculated using the absolute power values averaged for all electrodes as follows: [(delta + theta)/(beta)]. Recently it has been suggested that metrics describing network interactions of large-scale inter-areal synchronization between brain oscillations could improve classification accuracy in iRBD patients.^30^ Accordingly, overall functional connectivity for each frequency band was extracted by averaging the wPLI values of all 1770 electrode pairs.^8,31^ Furthermore, Shannon entropy (SE) was defined with 10 bins of amplitude values.^32^ In total, 15 EEG features were calculated for analysis (Supplementary Table 1). All spectral analyses were performed using the FieldTrip toolbox version 20200607.^33^

#### Modeling process (1) - prediction of phenoconversion time

Eighty percent of the overall dataset was assigned as the training set, and the remaining 20% of the data were designated as the testing set. All iRBD patient data, which were divided into iRBD patients not phenoconverted at follow-up (iRBD-NC) and iRBD patients phenoconverted at follow-up (iRBD-C), were used in this survival prediction analysis. All duration information were calculated by the difference between the EEG acquisition date and the last visit date of the patient. To identify the most relevant features for predicting phenoconversion time, a two-step feature selection process was used. First, univariable Cox proportional hazard (CPH) regression was performed to identify features with a p value less than 0.1. Next, backward multivariable CPH regression was performed to eliminate features with a p value greater than 0.1. To address the imbalanced nature of the data, with a larger number of nonconverter samples, the synthetic minority oversampling technique (SMOTE) was applied to the training set.^34^ CPH, Weibull-accelerated failure time (wAFT) and random survival forest (RSF) models were used to train and test these data. To evaluate the models, stratified group 5-fold cross-validation was implemented for internal validation. Harrel’s concordance index (C-index) and the integrated Brier score (IBS) were used to evaluate the performance of survival prediction analysis.^35,36^ The C-index is a commonly used metric in survival analysis that measures the ability of the model to correctly rank the survival times of patients.. The IBS, on the other hand, is a measure of the overall accuracy of the model’s predictions, considering both the predicted survival times and the observed survival times.. For all models, hyperparameter optimization was performed using the training set while the testing set was used for model performance. The final model was fitted with the augmented and resampled total dataset by using the best prediction model. As there were no MSA patients in the external validation set, we additionally fitted the model excluding the MSA patients. Permutation feature importance was employed to represent the importance of each variable in the model. All analyses were conducted using Python 3.8.5 (scikit-learn: v.1.1.1; lifelines: v.0.27.0; scikit-survival: v.0.18.0; hyperopt: v.0.2.7).

#### Modeling process (2) - prediction of phenoconversion subtype

Only iRBD-C data were used for subtype prediction analysis to classify subtypes of phenoconversion. A training set made up of 80% of the dataset and a testing set using the remaining 20% were assigned. Following a previous study, we classified phenoconversion into two subtypes according to the first presenting symptom: motor-first subtype, which includes PD and MSA, and cognition-first subtype, which includes DLB.^13^ Recursive feature elimination (RFE) was applied using multiple models to select the most relevant and predictive features for subtype prediction. The features selected by RFE using XGBoost showed the best performance and were therefore used for the models. The other models were trained and tested on the selected features. Due to data imbalance, the SMOTE was applied in the training set. The data were trained and tested using the XGBoost, random forest (RF), logistic regression (LR) with elastic net regularization and k-nearest neighbor (KNN) models. Hyperparameter optimization was performed using the training set for all models while evaluating by the testing set. A 10-fold cross-validation was performed 10 times for internal validation to obtain a robust estimate of the performance of the models. The area under the receiver operating characteristic curve (AUC), accuracy, precision, recall and F1 score were utilized to evaluate the performance of the subtype prediction analysis. The best prediction model was used to fit the final model to the augmented and resampled entire dataset. For the same reason described above for survival prediction, a dataset without data from MSA patients was additionally analyzed. Classification into PD, MSA and DLB was also performed. Python 3.8.5 was used to conduct each analysis (scikit-learn: v.1.1.1; xgboost: v.0.90; hyperopt: v.0.2.7).

### Statistical analysis

All data are shown as the mean ± standard deviation [range]. The Kolmogorov□Smirnov test was used to test the normality of all variables before analysis. Independent sample t tests were employed to evaluate differences in continuous data. Categorical data were analyzed with Fisher’s exact test. Nonnormally distributed variables were compared using the MannDWhitney U test. Survival curves were plotted using the Kaplan□Meier method. The log-rank test was used to compare survival distributions between our dataset and the external validation dataset. The Restricted Mean Survival Time (RMST) is obtained to estimate the average survival time up to 5.8 years, which is the last observed event time in our dataset.^37^ The significance threshold was set to 0.05. All statistical evaluations were performed with Python 3.8.5 using the SciPy library (scipy: v.1.5.2).

## Results

### Participant characteristics

A total of 236 iRBD patients were included in the internal validation dataset. During a mean follow-up duration of 3.5 years [range: 0.9–8.6 years], 31 patients converted to overt neurodegenerative diseases, and 205 remained in an isolated state of RBD (Figure 1A). The mean time to phenoconversion was 2.66 ± 1.48 years. 83 patients with no baseline EEG data, 1 patient who phenoconverted within 6 months after EEG acquisition and 10 patients whose data were recorded with another EEG system were excluded from further analysis. Of the remaining 142 patients, 27 patients phenoconverted during follow-up (13 to PD, 8 to DLB, and 6 to MSA). All MSA patients were of the cerebellar type (MSA-C).

**Figure 1.**
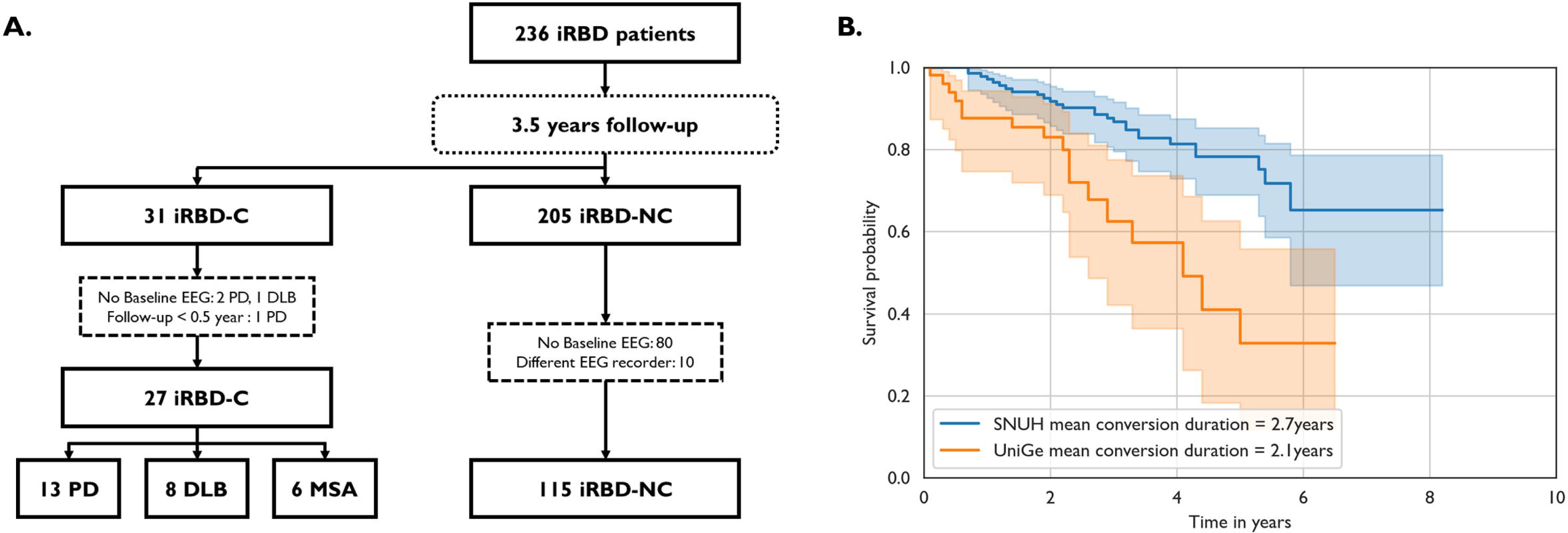
Flowchart and survival curve. (A) Flowchart. (B) Survival curves of Seoul National University Hospital and University of Genoa. Abbreviations: iRBD, isolated REM sleep behavior disorder; iRBD-C, iRBD converters; iRBD-NC, iRBD nonconverters; PD, Parkinson’s disease; DLB, dementia with Lewy bodies; MSA, multiple system atrophy; SNUH, Seoul National University Hospital; UniGe, University of Genoa.

There were no significant differences in sex, RBDQ-KR, K-MMSE, KVSS, SCOPA-AUT, ESS and PSQI between the iRBD-NC and iRBD-C groups. However, the patients in the iRBD-C group were older, had lower education levels, lower MoCA-K scores and higher MDS-UPDRS-III scores (Table 1). When comparing motor- and cognition-first subtypes, the cognition-first group was older and had lower baseline K-MMSE and MoCA-K scores (Supplementary Table 2).

**Table 1.**
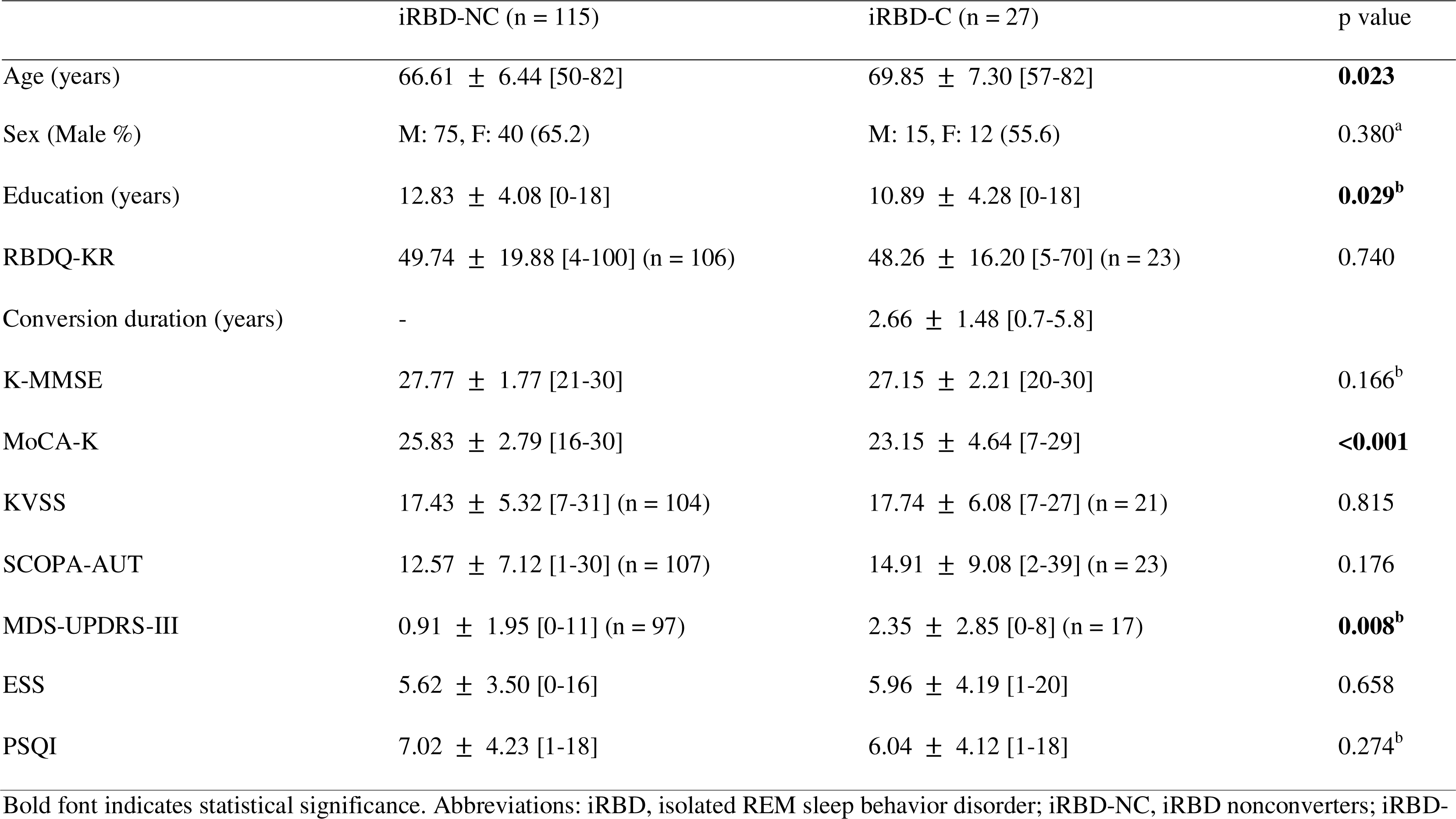
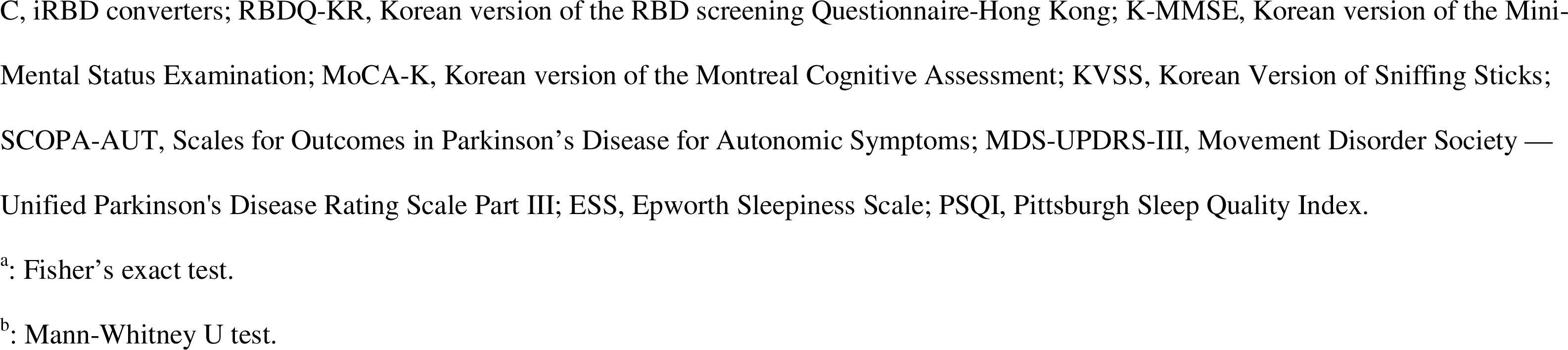
Participant characteristics of iRBD patients who further converted or not from Seoul National University Hospital.

|  | iRBD-NC (n = 115) | iRBD-C (n = 27) | p value |
| --- | --- | --- | --- |
| Age (years) | 66.61 ± 6.44 [50-82] | 69.85 ± 7.30 [57-82] | **0.023** |
| Sex (Male %) | M: 75, F: 40 (65.2) | M: 15, F: 12 (55.6) | 0.380<sup>a</sup> |
| Education (years) | 12.83 ± 4.08 [0-18] | 10.89 ± 4.28 [0-18] | **0.029<sup>b</sup>** |
| RBDQ-KR | 49.74 ± 19.88 [4-100] (n = 106) | 48.26 ± 16.20 [5-70] (n = 23) | 0.740 |
| Conversion duration (years) | - | 2.66 ± 1.48 [0.7-5.8] |  |
| K-MMSE | 27.77 ± 1.77 [21-30] | 27.15 ± 2.21 [20-30] | 0.166<sup>b</sup> |
| MoCA-K | 25.83 ± 2.79 [16-30] | 23.15 ± 4.64 [7-29] | **<0.001** |
| KVSS | 17.43 ± 5.32 [7-31] (n = 104) | 17.74 ± 6.08 [7-27] (n = 21) | 0.815 |
| SCOPA-AUT | 12.57 ± 7.12 [1-30] (n = 107) | 14.91 ± 9.08 [2-39] (n = 23) | 0.176 |
| MDS-UPDRS-III | 0.91 ± 1.95 [0-11] (n = 97) | 2.35 ± 2.85 [0-8] (n = 17) | **0.008<sup>b</sup>** |
| ESS | 5.62 ± 3.50 [0-16] | 5.96 ± 4.19 [1-20] | 0.658 |
| PSQI | 7.02 ± 4.23 [1-18] | 6.04 ± 4.12 [1-18] | 0.274<sup>b</sup> |

Bold font indicates statistical significance. Abbreviations: iRBD, isolated REM sleep behavior disorder; iRBD-NC, iRBD nonconverters; iRBD-
C, iRBD converters; RBDQ-KR, Korean version of the RBD screening Questionnaire-Hong Kong; K-MMSE, Korean version of the Mini-Mental Status Examination; MoCA-K, Korean version of the Montreal Cognitive Assessment; KVSS, Korean Version of Sniffing Sticks; SCOPA-AUT, Scales for Outcomes in Parkinson's Disease for Autonomic Symptoms; MDS-UPDRS-III, Movement Disorder Society — Unified Parkinson's Disease Rating Scale Part III; ESS, Epworth Sleepiness Scale; PSQI, Pittsburgh Sleep Quality Index.
<sup>a</sup>: Fisher's exact test.
<sup>b</sup>: Mann-Whitney U test.

The external validation dataset included 62 iRBD patients who were followed up for 2.17 ± 1.53 years (Supplementary Table 3). Seven patients were excluded: 5 because of poor data quality, 1 because the data does not have conversion date information and 1 because the data were recorded after phenoconversion. Seventeen of the iRBD patients in the external validation dataset were phenoconverted during follow-up (7 to PD and 10 to DLB). Compared to our dataset, patients in the external validation dataset were older, were more likely to be male and had lower MMSE scores (Supplementary Table 4). In addition, the log-rank test showed that the two datasets showed significantly different survival rates (Figure 1B, *p* < 0.005). The RMST of our dataset was 5.067 years and the RMST of each patient was calculated from the survival curve. In addition, a positive correlation was shown between the RMST and time to conversion for the converted patients (Pearson correlation r = 0.525, *p* = 0.005, Supplementary Figure 1).

### Phenoconversion time prediction

Delta wPLI was excluded through univariable CPH regression, and DOF, relative delta power, relative beta power and SE were further excluded through multivariable CPH regression. Finally, 10 features were included in this survival prediction analysis. The power spectral densities of iRBD-NC and iRBD-C are shown in Figure 2B.

**Figure 2.**
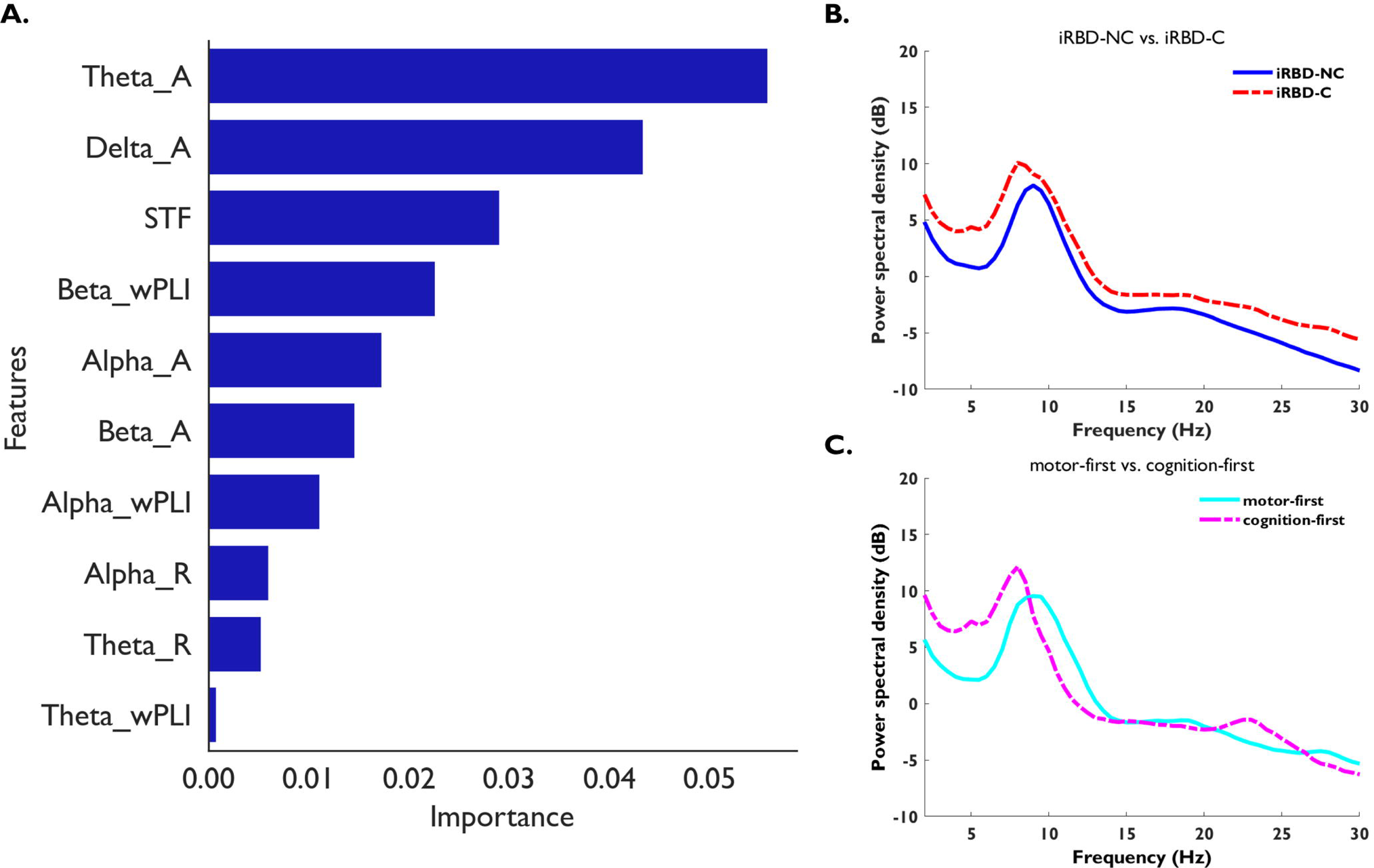
(A) Feature importance of random survival forest model and (B) comparison of power spectral densities between the iRBD-NC and iRBD-C groups, and (C) comparison of motor-first and cognition-first presentations. Abbreviations: _A, absolute power; STF, slow-to-fast power ratio; wPLI, weighted phase lag index; _R, relative power; SE, Shannon entropy; iRBD, isolated REM sleep behavior disorder; iRBD-C, converters to a neurodegenerative disorder from iRBD; iRBD-NC, nonconverters from iRBD.

We compared the three survival analysis methods using our dataset (Supplementary Table 5). For the internal validation using 5-fold cross-validation, the RSF model was the best, with an IBS of 0.114 and a C-index of 0.775. The five most important features of RSF were absolute theta power, absolute delta power, STF, beta wPLI and absolute alpha power (Figure 2A and Supplementary Table 6). The iRBD-C group showed higher absolute delta power and absolute theta power but also higher alpha power than the iRBD-NC group. For the external validation dataset, the RSF model showed an IBS of 0.128 and a C-index of 0.561.

Additionally, model results excluding data from MSA patients are listed in Supplementary Table 7.

### Phenoconversion subtype prediction

Through recursive feature elimination, eight features were excluded due to their low feature importance. As a result, seven features were used in this subtype prediction analysis. The selected features in subtype prediction were DOF, STF, absolute theta power, absolute beta power, relative beta power, beta wPLI and SE (Supplementary Table 8). The power spectral densities of the motor- and cognition-first subtypes are shown in Figure 2C.

The scores for internal validation of motor- and cognition-first are shown in Table 2. For internal validation, the KNN model’s performance was the best among the models, with an AUC of 0.901, accuracy of 0.704, precision of 0.500, recall of 0.875 and F1 of 0.636 (Figure 3). External validation of the KNN model resulted in an AUC of 0.536, accuracy of 0.527, precision of 0.304, recall of 0.412 and F1 of 0.300.

**Figure 3.**
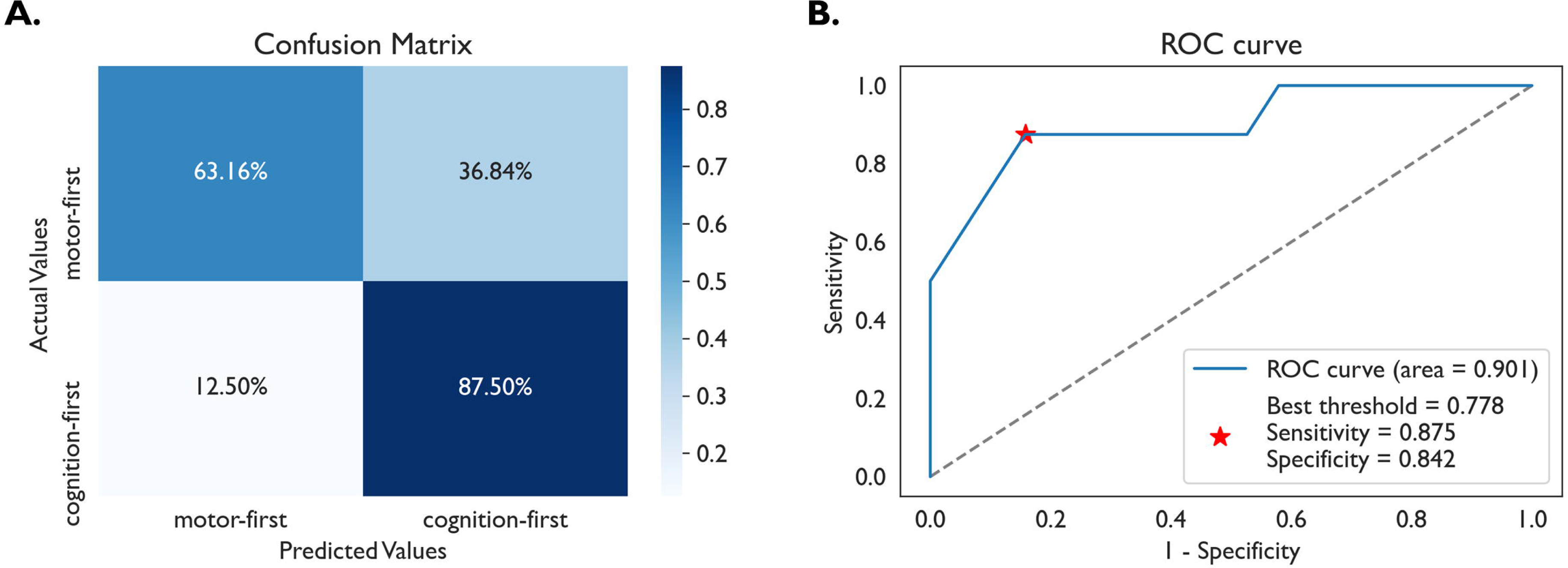
K-nearest neighbor model prediction results. These results are obtained by internal validation using repeated 10-fold cross-validation. (A) Confusion matrix. (B) Receiver operating characteristic curve. Abbreviations: ROC, receiver operating characteristic curve.

**Table 2.** Subtype prediction results.

|  | AUC | Accuracy | Precision | Recall | F1 |
| --- | --- | --- | --- | --- | --- |
| XGBoost | 0.717 | 0.741 | 0.556 | 0.625 | 0.588 |
| RF | 0.737 | 0.704 | 0.500 | 0.750 | 0.600 |
| LR | 0.770 | 0.778 | 0.667 | 0.500 | 0.571 |
| KNN | 0.901 | 0.704 | 0.500 | 0.875 | 0.636 |

Abbreviations: XGBoost, extreme gradient boosting; RF, random forest; LR, logistic regression; KNN, K-nearest neighbor; AUC, area under the receiver operating characteristic curve.

In addition, evaluation results without data from MSA patients and classification into PD, MSA and DLB are shown in Supplementary Tables 9 and 10, respectively. Example plots using both phenoconversion time and subtype prediction models are shown in Supplementary Figures 2 and 3.

## Discussion

In this study, we aimed to predict phenoconversion time and subtype in iRBD patients using resting-state EEG features collected at baseline. Our models, which were based on machine learning algorithms, showed promising results in predicting phenoconversion time and subtype. The RSF model showed acceptable performance in predicting phenoconversion time, while the KNN model was able to predict the conversion subtype (motor-first or cognition-first) with good AUC. Our models may provide a practical solution for predicting individualized phenoconversion time and subtype in iRBD patients. These predictions are important for better management of the disease and to help patients to be better prepared for their future.

Two previous studies have attempted to predict phenoconversion from iRBD patients using EEG. In the first, researchers used EEG slowing features to predict neurodegeneration in iRBD patients.^13^ The focus was to classify iRBD whether patients would phenoconvert without considering phenoconversion time or specific subtypes. Later, the same research group used deep learning techniques with EEG spectrograms recorded from iRBD patients to differentiate them from healthy controls.^38^ In contrast, our study aimed to predict not only the phenoconversion time but also the conversion subtype of iRBD patients. We predicted phenoconversion time in patients with iRBD using only baseline EEG features. Absolute theta power, absolute delta power, beta wPLI, STF and absolute alpha power were the most important features for phenoconversion time prediction. In previous studies, it was shown that the absolute EEG power of recordings during both sleep and resting-state was significantly different not only between iRBD patients and controls but also between iRBD patients who converted to neurodegenerative diseases and those who had not yet converted.^12,13,38–41^ In particular, the increases in absolute theta power and delta power were prominent in converted patients. Higher low-frequency power and lower high-frequency power, called EEG slowing, have already been shown in iRBD patients by various neurodegenerative studies.^13,39,42–44^ Therefore, EEG slowing is known to be common in iRBD patients, particularly in those who convert to neurodegenerative diseases. Our results demonstrate that EEG features can be applied as biomarkers for predicting phenoconversion time in iRBD patients.

Phenoconversion subtype prediction from iRBD patients was also feasible using baseline EEG features. EEG differences between the motor- and cognition-first subtypes had been shown in a previous study. The cognition-first subtype (DLB patients) showed increased delta and theta power, higher STF and lower DOF.^45,46^ Indeed, the selected features for subtype prediction in our study were DOF, STF, absolute theta and beta power, which are consistent with previous studies. Additionally, EEG slowing is correlated with cognitive impairment.^47^ In previous studies comparing the motor- and cognition-first subtypes, the main difference at baseline was cognitive function, which was significantly decreased in the cognition-first subtype.^2,48,49^

Exclusion of MSA patients slightly improved the performance of the phenoconversion time prediction model (Supplementary Table 7). The reduced heterogeneity of the sample may have made it easier for the model to identify the relevant features for phenoconversion time prediction. However, it reduced the performance of the KNN model for phenoconversion subtype prediction. The decrease in the number of converters in the dataset from 27 to 21 following the exclusion of 6 MSA patients could have contributed to a significant decrease in the model performance.

It is notable that the performance of external validation was not as good as expected for both survival prediction and subtype prediction. Significantly lower performance for the external validation dataset indicates that our model is overfitted to our dataset. Although we took measures to prevent overfitting by conducting cross-validation and adjusting parameters known to affect overfitting, we could not fully escape overfitting. Compared to the external validation dataset, our dataset showed a higher proportion of females, younger age, lower MDS-UPDRS-III scores and more MSA converted patients. Moreover, lower number of subjects in the external validation dataset may have also affected the performance of our model. These significant differences between the two cohorts might have contributed to the poor performance in the external validation in this study.

Studies conducted in cohorts from Asian countries have found a slower phenoconversion rate than those from European and American cohorts, suggesting ethnic differences in the prognosis of iRBD.^50,51^ If this is true, ethnic differences should be taken into consideration in the prediction model. Further study is mandatory to confirm any ethnic and/or regional differences in the prognosis of iRBD in the future.

There are a few limitations to note. First, age and cognitive function scores, which may affect EEG findings were not accounted for.^52,53^ Second, due to the small sample size, we were forced to apply data augmentation. The number of iRBD patients in this study was 143, which was relatively small for the use of machine learning methods. However, as many studies have used EEG sliding window data augmentation, this method of data augmentation is likely to be reliable enough to achieve the goal of our study.^54–56^

In conclusion, we were able to create a useful RSF model and KNN model for predicting the time of phenoconversion and its subtype, respectively, in iRBD patients simply using resting-state EEG features at baseline. We believe our prediction model and method contribute to opening new horizons in the management and counseling of iRBD patients. Furthermore, our model can be implemented in clinical EEG machines or can be developed as a stand-alone device that can be used in outpatient clinics. A future multicenter study with a larger number of patients is needed to elucidate the predictive value of baseline EEG features.

## Acknowledgment

This work was supported by the Brain Research Program through the National Research Foundation of Korea (NRF) funded by the Ministry of Science, ICT & Future Planning (2017M3C7A1029688) and the National Research Foundation of Korea (NRF) grant funded by the Korean government (MSIP) (2017R1A2B2012280). This work was partially supported by a grant from the Italian Ministry of Health to IRCCS Ospedale Policlinico San Martino (Fondi per la Ricerca Corrente 2019/2020, and Italian Neuroscience network (RIN)). This work was developed within the framework of the DINOGMI Department of Excellence of MIUR 2018-2022 (legge 232 del 2016). This work was carried out within the framework of the project "RAISE - Robotics and AI for Socio-economic Empowerment” and has been supported by European Union - NextGenerationEU.

## Data availability statement

The data that support the findings of this study are available from the corresponding author upon reasonable request.

## Author’s roles

El Jeong: Methodology, Formal analysis, Software, Writing – original draft preparation

Yong Woo Shin: Methodology, Formal analysis, Software, Writing – original draft preparation

Jung-Ick Byun: Supervision, Data curation, Validation, Writing – review & editing

Jun-Sang Sunwoo: Data curation, Validation, Investigation

Monica Roascio: Data curation, Validation, Investigation, Writing – review & editing

Pietro Mattioli: Data curation, Validation, Investigation

Laura Giorgetti: Data curation, Validation, Investigation

Francesco Famà: Data curation, Validation, Investigation

Gabriele Arnulfo: Data curation, Validation, Investigation, Writing – review & editing

Dario Arnaldi: Data curation, Validation, Investigation, Writing – review & editing

Han-Joon Kim: Resources, Validation, Investigation

Ki-Young Jung: Supervision, Writing – review & editing, Project administration, Funding acquisition

## Notes

**Conflicts of interest:** Nothing to report.

**Funding:** This work was supported by the Brain Research Program through the National Research Foundation of Korea (NRF) funded by the Ministry of Science, ICT & Future Planning (2017M3C7A1029688) and the National Research Foundation of Korea (NRF) grant funded by the Korean government (MSIP) (2017R1A2B2012280). This work was partially supported by a grant from the Italian Ministry of Health to IRCCS Ospedale Policlinico San Martino (Fondi per la Ricerca Corrente 2019/2020, and Italian Neuroscience network (RIN)). This work was developed within the framework of the DINOGMI Department of Excellence of MIUR 2018-2022 (legge 232 del 2016). This work was carried out within the framework of the project "RAISE - Robotics and AI for Socio-economic Empowerment” and has been supported by European Union - NextGenerationEU.

### Competing Interest Statement

The authors have declared no competing interest.

### Author Declarations

IRB of Seoul National University Hospital (IRB Number 1406-100-589) gave ethical approval for this work. IRB of University Neurology Clinics at Policlinico San Martino in Genoa (IRB Number 703) gave ethical approval for this work.

