## Supplementary figures and images for "EEG-based Machine Learning Models for the Prediction of Phenoconversion Time and Subtype in iRBD"

### Supplementary figure 1

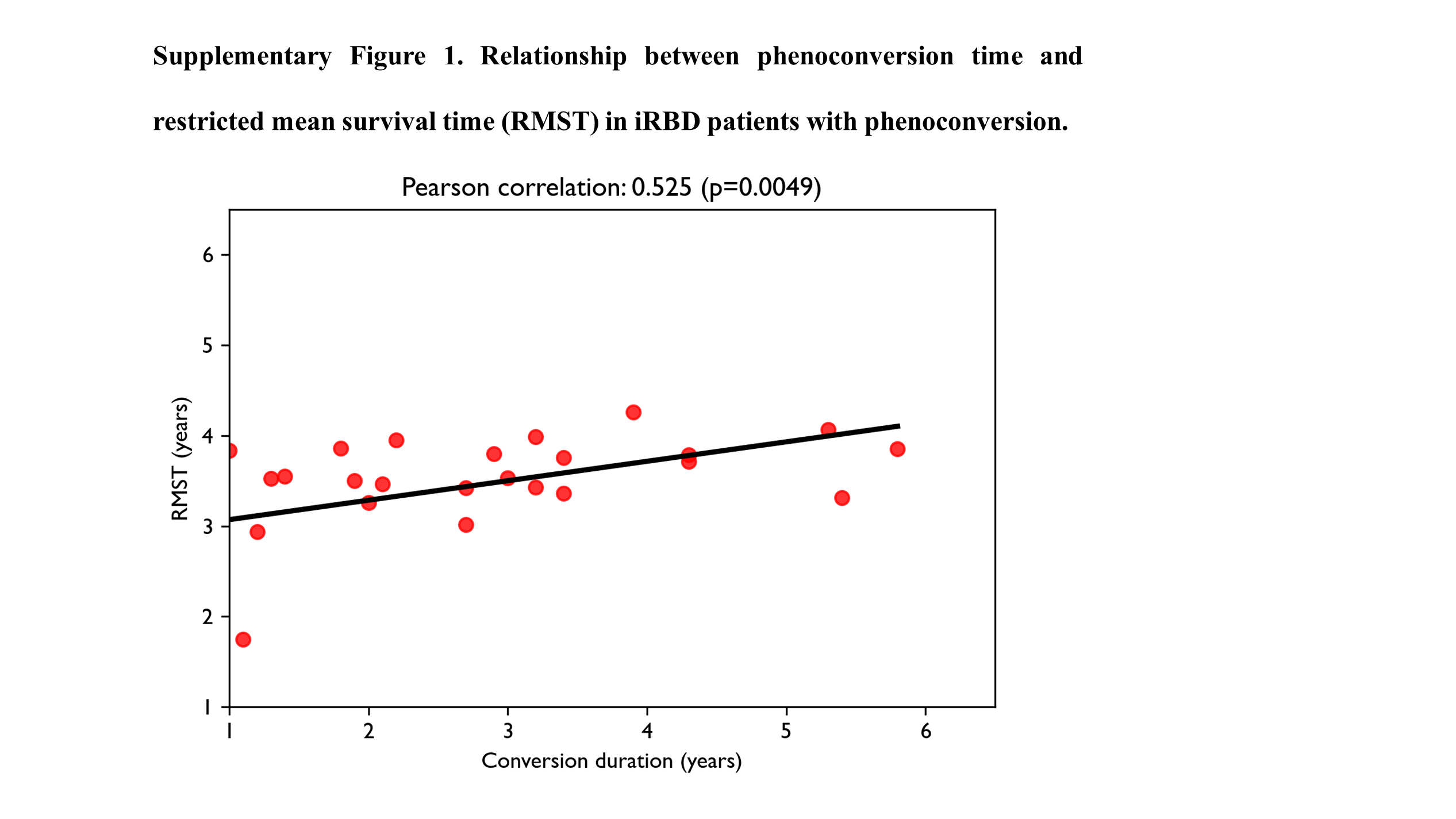

### Supplementary figure 2

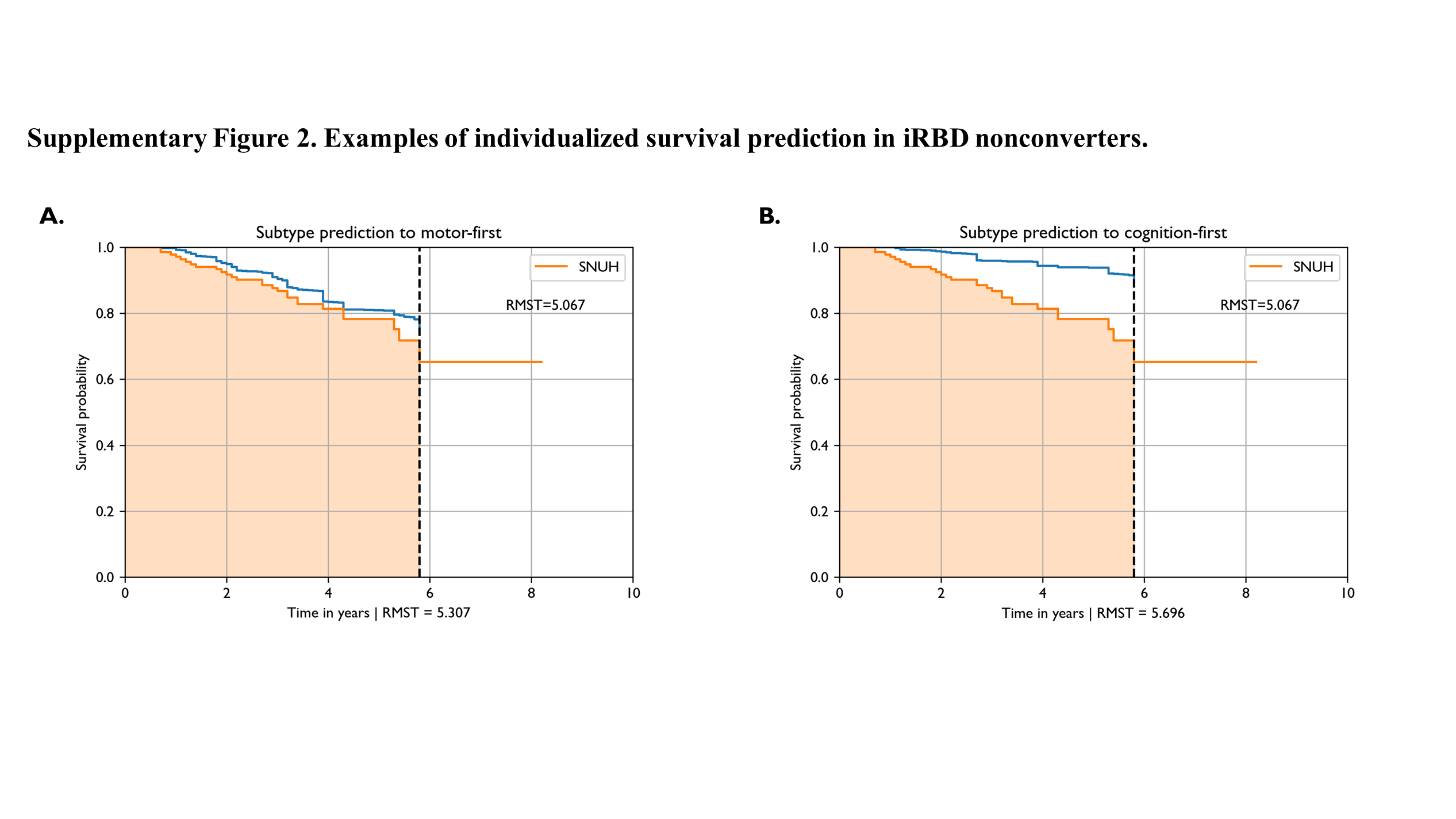

### Supplementary figure 3

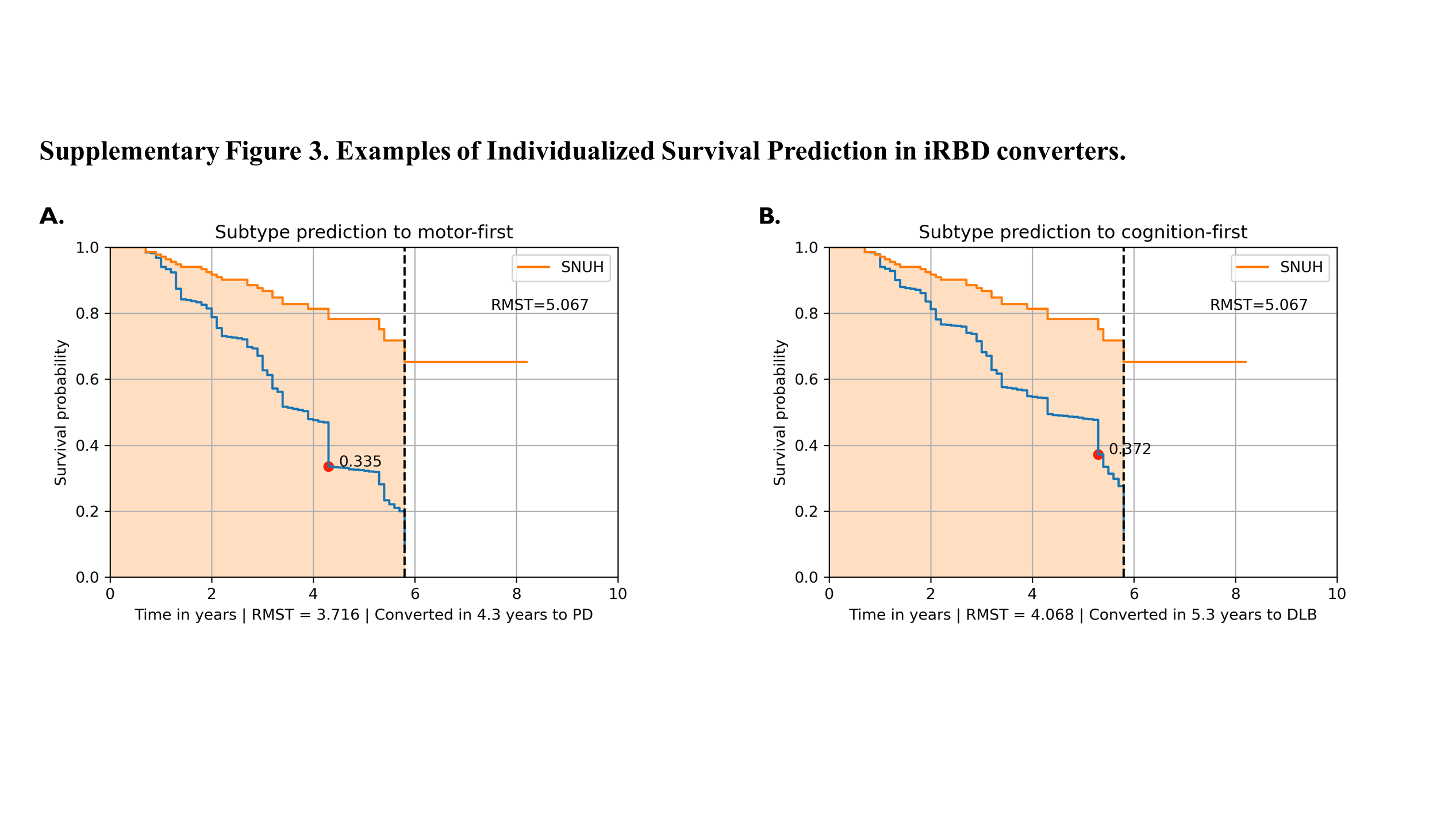
