## Supplementary tables for "EEG-based Machine Learning Models for the Prediction of Phenoconversion Time and Subtype in iRBD"

### **Supplementary Table 1. EEG features**

| Categories | Features |
| --- | --- |
| Frequency-domain | Absolute delta power: 2-3.5 Hz |
|  | Absolute theta power: 4-7.5 Hz |
|  | Absolute alpha power: 8-12.5 Hz |
|  | Absolute beta power: 13-30 Hz |
|  | Relative delta power: 2-3.5 Hz |
|  | Relative theta power: 4-7.5 Hz |
|  | Relative alpha power: 8-12.5 Hz |
|  | Relative beta power: 13-30 Hz |
|  | DOF: occipital peak frequency |
|  | Slow-to-fast power ratio: (delta + theta)/beta |
| Functional connectivity | Delta wPLI value |
|  | Theta wPLI value |
|  | Alpha wPLI value |
|  | Beta wPLI value |
| Entropy | Shannon entropy: 10 bins of amplitude |

Abbreviations: DOF, dominant occipital frequency; wPLI, weighted phase lag index.

### **Supplementary Table 2. Participant characteristics of motor- and cognition-first subtypes from Seoul National University Hospital**

|  | Motor-first (n = 19) | Cognition-first (n = 8) | p value |
| --- | --- | --- | --- |
| Age (years) | 67.79 $\pm$ 6.57 [57-80] | 74.75 $\pm$ 6.92 [61-82] | **0.020** |
| Sex (Male %) | M: 10, F: 9 (52.6) | M: 5, F: 3 (62.5) | 0.696^a^ |
| Education (years) | 11.37 $\pm$ 3.86 [6-16] | 9.75 $\pm$ 5.26 [0-18] | 0.380 |
| RBDQ-KR | 49.29 $\pm$ 11.86 [19-66] (n = 17) | 45.33 $\pm$ 26.27 [5-70] (n = 6) | 0.618 |
| Conversion duration (years) | 2.89 $\pm$ 1.45 [0.7-5.8] | 2.11 $\pm$ 1.51 [0.7-5.3] | 0.219 |
| K-MMSE | 27.79 $\pm$ 1.65 [24-30] | 25.63 $\pm$ 2.72 [20-28] | **0.017** |
| MoCA-K | 24.63 $\pm$ 3.25 [19-29] | 19.63 $\pm$ 5.71 [7-24] | **0.008** |
| KVSS | 18.57 $\pm$ 5.75 [10-27] (n = 15) | 15.67 $\pm$ 6.92 [7-27] (n = 6) | 0.336 |
| SCOPA-AUT | 14.44 $\pm$ 9.77 [2-39] (n = 18) | 16.60 $\pm$ 6.58 [9-23] (n = 5) | 0.649 |
| MDS-UPDRS-III | 2.40 $\pm$ 3.04 [0-8] (n = 15) | 2.00 $\pm$ 0.00 [2-2] (n = 2) | 0.697^b^ |
| ESS | 6.11 $\pm$ 4.50 [1-20] | 5.63 $\pm$ 3.62 [2-13] | 0.792 |
| PSQI | 6.42 $\pm$ 3.93 [2-18] | 5.13 $\pm$ 4.67 [1-16] | 0.466 |

Bold font indicates statistical significance. Abbreviations: RBDQ-KR, Korean version of the RBD screening Questionnaire-Hong Kong; K-MMSE, Korean version of the Mini-Mental Status Examination; MoCA-K, Korean version of the Montreal Cognitive Assessment; KVSS, Korean Version of Sniffing Sticks; SCOPA-AUT, Scales for Outcomes in Parkinson’s Disease for Autonomic Symptoms; MDS-UPDRS-III, Movement Disorder Society — Unified Parkinson's Disease Rating Scale Part III; ESS, Epworth Sleepiness Scale; PSQI, Pittsburgh Sleep Quality Index.

^a^: Fisher’s exact test.

^b^: Mann-Whitney U test.

### **Supplementary Table 3. Participant characteristics of iRBD patients who further converted or not from University of Genoa**

|  | iRBD-NC (n = 38) | iRBD-C (n = 17) | p value |
| --- | --- | --- | --- |
| Age (years) | 69.71 $\pm$ 6.70 [56-81] | 70.94 $\pm$ 6.55 [60-84] | 0.529 |
| Sex (Male %) | M: 33, F: 5 (86.8) | M: 13, F: 4 (76.5) | 0.435**^a^** |
| Conversion duration (years) | - | 2.05 $\pm$ 1.49 [0.1-5.0]  (7 PD, 10 DLB) |  |
| MMSE | 28.53 $\pm$ 1.33 [25-30] | 27.06 $\pm$ 3.40 [17-30] | 0.161**^b^** |
| MDS-UPDRS-III | 1.53 $\pm$ 3.72 [0-19] (n = 32) | 1.88 $\pm$ 2.32 [0-8] | 0.101**^b^** |

Abbreviations: iRBD, isolated REM sleep behavior disorder; iRBD-NC, iRBD nonconverters; iRBD-C, iRBD converters; PD, Parkinson’s disease; DLB, dementia with Lewy bodies; MMSE, Mini-Mental Status Examination; MDS-UPDRS-III, Movement Disorder Society — Unified Parkinson's Disease Rating Scale Part III.

^a^: Fisher’s exact test.

^b^: Mann-Whitney U test.

### **Supplementary Table 4. Participant characteristics of iRBD patients from Seoul National University Hospital and University of Genoa**

|  | SNUH (n = 142) | UniGe (n = 55) | p value |
| --- | --- | --- | --- |
| Age (years) | 67.79 $\pm$ 6.87 [50-82] | 69.33 $\pm$ 6.28 [57-84] | **0.008** |
| Sex (Male %) | M: 90, F: 52 (63.4) | M: 46, F: 9 (83.6) | **0.006^a^** |
| Conversion duration (years) | 2.66 ± 1.48 [0.7-5.8] (n = 27) | 2.05 $\pm$ 1.49 [0.1-5.0] (n = 17) | 0.199 |
| MMSE | 27.81 $\pm$ 1.85 [20-30] | 27.50 $\pm$ 2.68 [17-30] | **0.031^b^** |
| MDS-UPDRS-III | 1.10 $\pm$ 2.49 [0-19] (n = 114) | 2.27 $\pm$ 2.65 [0-8] (n = 49) | 0.312^b^ |

Bold font indicates statistical significance. Abbreviations: iRBD, isolated REM sleep behavior disorder; SNUH, Seoul National University Hospital; UniGe, University of Genoa; MMSE, Mini-Mental Status Examination; MDS-UPDRS-III, Movement Disorder Society — Unified Parkinson's Disease Rating Scale Part III.

^a^: Fisher’s exact test.

^b^: Mann-Whitney U test.

### **Supplementary Table 5. Survival prediction results**

|  | IBS | C-index |
| --- | --- | --- |
| CPH | 0.161 | 0.767 |
| wAFT | 0.170 | 0.764 |
| RSF | 0.114 | 0.775 |

Abbreviations: IBS, integrated Brier score; C-index, concordance index; CPH, Cox proportional hazard; wAFT, Weibull-accelerated failure time; RSF, random survival forest.

### **Supplementary Table 6. Five most important features of random survival forest model for iRBD patients who further converted or not**

|  | iRBD-NC (n = 115) | iRBD-C (n = 27) | p value |
| --- | --- | --- | --- |
| Absolute theta power ($dB)$ | 2.72 $\pm$ 3.11 | 6.63 $\pm$ 3.76 | **<0.001** |
| Absolute delta power ($dB)$ | 4.56 $\pm$ 2.25 | 6.69 $\pm$ 2.93 | **<0.001** |
| STF | 3.66 $\pm$ 3.96 | 2.42 $\pm$ 0.66 | 0.636^a^ |
| Beta wPLI | 0.31 $\pm$ 0.06 | 0.27 $\pm$ 0.06 | **0.001** |
| Absolute alpha power ($dB)$ | 6.03 $\pm$ 4.44 | 9.16 $\pm$ 3.14 | **<0.001** |

Bold font indicates statistical significance. Abbreviations: iRBD, isolated REM sleep behavior disorder; iRBD-NC, iRBD nonconverters; iRBD-C, iRBD converters; STF, slow-to-fast power ratio; wPLI, weighted phase lag index.

p value: age and sex adjusted by analysis of covariance.

^a^: Mann-Whitney U test.

### **Supplementary Table 7. Survival prediction results without data from multiple system atrophy patients**

|  | IBS | C-index |
| --- | --- | --- |
| CPH | 0.129 | 0.815 |
| wAFT | 0.133 | 0.814 |
| RSF | 0.102 | 0.759 |

External validation: RSF IBS: 0.130; C-index: 0.543.

Abbreviations: IBS, integrated Brier score; C-index, concordance index; CPH, Cox proportional hazard; wAFT, Weibull-accelerated failure time; RSF, random survival forest.

### **Supplementary Table 8. Selected features of motor- and cognition-first subtypes in subtype prediction**

|  | Motor-first (n = 19) | Cognition-first (n = 8) | p value |
| --- | --- | --- | --- |
| DOF ($Hz)$ | 9.07 $\pm$ 0.87 | 7.75 $\pm$ 0.88 | **0.001** |
| STF | 2.19 $\pm$ 0.53 | 2.97 $\pm$ 0.64 | **0.003** |
| Absolute theta power ($dB)$ | 5.49 $\pm$ 3.05 | 9.36 $\pm$ 4.08 | **0.011** |
| Absolute beta power ($dB)$ | -0.49 $\pm$ 2.15 | -0.74 $\pm$ 3.36 | 0.817 |
| Relative beta power | 0.06 $\pm$ 0.04 | 0.04 $\pm$ 0.02 | 0.131 |
| Beta wPLI | 0.27 $\pm$ 0.06 | 0.27 $\pm$ 0.07 | 0.988 |
| Shannon entropy | 1.94 $\pm$ 0.04 | 1.91 $\pm$ 0.05 | 0.262 |

Bold font indicates statistical significance. Abbreviations: PD, Parkinson’s disease; MSA, multiple system atrophy; DLB, dementia with Lewy bodies; DOF, dominant occipital frequency; STF, Slow-to-fast power ratio; wPLI, weighted phase lag index.

p value: age and sex adjusted by analysis of covariance.

### **Supplementary Table 9. Subtype prediction results without data from multiple system atrophy patients**

|  | AUC | Accuracy | Precision | Recall | F1 |
| --- | --- | --- | --- | --- | --- |
| XGBoost | 0.615 | 0.619 | 0.500 | 0.625 | 0.556 |
| RF | 0.635 | 0.714 | 0.625 | 0.625 | 0.625 |
| LR | 0.663 | 0.619 | 0.500 | 0.500 | 0.500 |
| KNN | 0.615 | 0.667 | 0.571 | 0.500 | 0.533 |

External validation: LR AUC: 0.577; accuracy: 0.709; precision: 0.556; recall: 0.294; F1: 0.385.

Abbreviations: XGBoost, extreme gradient boosting; RF, random forest; LR, logistic regression; KNN, K-nearest neighbor; AUC, area under the receiver operating characteristic curve.

### **Supplementary Table 10. Subtype prediction results into Parkinson’s disease, multiple system atrophy and dementia with Lewy bodies**

|  | Accuracy |
| --- | --- |
| XGBoost | 0.407 |
| RF | 0.370 |
| LR | 0.222 |
| KNN | 0.407 |

External validation: KNN accuracy: 0.400.

Abbreviations: XGBoost, extreme gradient boosting; RF, random forest; LR, logistic regression; KNN, K-nearest neighbor.
