## Supplementary figure legends for "EEG-based Machine Learning Models for the Prediction of Phenoconversion Time and Subtype in iRBD"

**Supplementary Figure 1. Relationship between phenoconversion time and restricted mean survival time (RMST) in iRBD patients with phenoconversion.**

**Supplementary Figure 2. Examples of individualized survival prediction in iRBD nonconverters.**

(A) The predicted survival curve for a male patient in his 60s who has experienced dream enactment for 2 years. This patient has an initial MDS-UPDRS-III score of 0, a MoCA-K score of 25, and initially presented with depression and constipation, but no anosmia. The patient has been followed up for 6.9 years without phenoconversion. The prediction model suggests a motor-first subtype and an RMST value of 5.307, which is higher than the overall cohort's RMST of 5.067. (B) The predicted survival curve for a male patient in his 70s who began experiencing dream enactment for 3 years. This patient has an initial MDS-UPDRS-III score of 1 (right arm with fine postural tremor), a MoCA-K score of 27, no constipation and normal olfaction. The patient has been followed up for 6 years without phenoconversion, and at the 6-year mark, his MDS-UPDRS-III score was 2 and the MoCA-K score was 30. The prediction model indicates a motor-first subtype and an RMST value of 5.694, which is higher than the overall cohort's RMST of 5.067.

Abbreviations: iRBD, isolated rapid eye movement sleep behavior disorder; MDS-UPDRS-III, Movement Disorder Society — Unified Parkinson's Disease Rating Scale Part III; MoCA-K, Korean version of the Montreal Cognitive Assessment; RMST, restricted mean survival time.

**Supplementary Figure 3. Examples of Individualized Survival Prediction in iRBD converters.**

(A) The predicted survival curve for a female patient in her 60s with DEB that started 3 years prior and was diagnosed with iRBD. She initially reported subjective cognitive impairment, a K-MMSE score of 30, MoCA-K score of 25 and depressive mood (GDS score of 42). Her initial MDS-UPDRS-III score was 8, but she didn't meet degenerative disease criteria, and no constipation or anosmia was identified. After 4.6 years, she phenoconverted to Parkinson's disease, At the time of phenoconversion, her MoCA-K score was 21, and her MDS-UPDRS-III score was 9. with an RMST of 4.286, lower than the cohort's 5.067. The predicted subtype was motor-first, consistent with her clinical presentation. (B) The predicted survival curve for a male patient in his 70s who began experiencing DEB 5 years prior to his visit. He initially reported subjective cognitive impairment, with a K-MMSE score of 26, a CDR of 0.5 and a MoCA-K score of 24. His initial MDS-UPDRS-III score was 0, and he had constipation and hyposmia. Over time, his cognitive impairment worsened and after 6.4 years, he phenoconverted to DLB. At the time of phenoconversion, his MDS-UPDRS-III score became 8, and he had anosmia. His MoCA-K score had decreased to 21. The progression of his cognitive impairment and other symptoms ultimately led to a diagnosis of DLB. Her RMST was 4.068, which was lower than the cohort's 5.067. The predicted subtype was cognition-first, consistent with her clinical presentation.

Abbreviations: iRBD, isolated REM sleep behavior disorder; DEB, dream enactment behavior; K-MMSE, Korean version of the Mini-Mental Status Examination; MoCA-K, Korean version of the Montreal Cognitive Assessment; GDS, Geriatric Depression Scale; MDS-UPDRS-III, Movement Disorder Society — Unified Parkinson's Disease Rating Scale Part III; RMST, restricted mean survival time; CDR, Clinical Dementia Rating; DLB, dementia with Lewy bodies.
